# Vascular Remodelling in Idiopathic Nephrotic Syndrome with Administration of Angiotensin-Converting Enzyme Inhibitors and Angiotensin Receptor Blockers

**DOI:** 10.64898/2026.10.09.26365110

**Authors:** Syafruddin Haris, Herlina Dimiati, Kartini Hasballah, Sudung O Pardede

## Abstract

Pediatric nephrotic syndrome (NS) significantly increases cardiovascular risk, primarily through activation of the renin-angiotensin-aldosterone system (RAAS) and chronic inflammation. This study aimed to evaluate the relationship between serum aldosterone and interleukin 6 (IL-6) levels with carotid intima-media thickness (CIMT) in children with steroid-sensitive (SSNS) and steroid-resistant NS (SRNS), before and after treatment with angiotensin-converting enzyme inhibitors (ACEI) and angiotensin receptor blockers (ARB). This prospective, single-arm pretest–posttest study included 32 children aged 2–18 years with idiopathic NS at Dr. Zainoel Abidin Hospital, Banda Aceh, Indonesia. Participants received lisinopril and valsartan for 12 weeks. Serum aldosterone was quantified by liquid chromatography-tandem mass spectrometry, IL-6 by electrochemiluminescence immunoassay, and CIMT by high-resolution B-mode Doppler ultrasound. Data were analyzed using SPSS version 26.0, including Shapiro-Wilk normality testing, correlation analyses, and multivariate linear regression, with statistical significance set at p<0.05. CIMT decreased significantly after treatment (0.13±0.04 cm to 0.06±0.01 cm; p<0.001). Serum aldosterone and IL-6 did not change significantly. Multivariate regression identified steroid resistance as associated with CIMT change, while aldosterone and IL-6 showed no significant independent effects.

## INTRODUCTION

Nephrotic syndrome (NS) is the leading cause of renal impairment in children, characterized by massive proteinuria, hypoalbuminemia, dyslipidemia, and generalized edema [1]. NS significantly increases morbidity and mortality, mainly from cardiovascular complications [1, 2]. Children with NS exhibit elevated cardiovascular risk attributed to hyperlipidemia, increased thrombogenesis, and endothelial dysfunction [2, 3], compounded by steroid toxicity, persistent proteinuria, oxidative stress, recurrent infections, thromboembolism, and nephrotoxic treatments such as calcineurin inhibitors [3, 4].

NS is a major cause of pediatric chronic kidney disease (CKD), defined by structural or functional kidney abnormalities lasting over three months [5]. Between 85–90% of children with idiopathic NS respond to corticosteroid therapy and are classified as having steroid-sensitive nephrotic syndrome (SSNS); however, 40–50% of these patients experience frequent relapses or develop steroid dependency [6, 7]. Approximately 10–15% exhibit steroid-resistance nephrotic syndrome (SRNS), which is associated with a higher risk of progression to end-stage renal disease (ESRD) and adverse cardiovascular outcomes [6-8].

Hypertension, a common and significant cardiovascular risk factor in pediatric NS and CKD, results from intrinsic renal dysfunction, fluid retention, treatment effects, and chronic renal fibrosis [9]. The renin-angiotensin-aldosterone system (RAAS) plays a critical role in regulating blood pressure, sodium balance, and renal hemodynamics [10]. Angiotensin II and aldosterone promote vasoconstriction, sodium retention, and vascular remodelling, which exacerbate hypertension and contribute to cardiovascular injury [10]. Accordingly, angiotensin-converting enzyme inhibitors (ACEIs) and angiotensin receptor blockers (ARBs), which inhibit RAAS, are essential for managing hypertension and preventing kidney damage in children with NS [11].

Chronic inflammation contributes substantially to cardiovascular complications in NS and CKD. Interleukin-6 (IL-6), a pro-inflammatory cytokine rapidly elevates in response to injury and infection, is associated with worsening kidney function and vascular damage [12-14]. IL-6 mediates endothelial dysfunction by reducing nitric oxide production and adiponectin expression, thereby accelerating atherosclerosis [15]. Carotid intima-media thickness (CIMT) measured by ultrasound is a sensitive, non-invasive indicator of early vascular alterations [16]. In children with NS, factors such as prolonged disease duration, frequent relapses, diastolic hypertension, and advancing age are correlated with increased CIMT, reflecting early vascular injury and heightened cardiovascular risk [17].

Although aldosterone and IL-6 independently contribute to vascular injury, their combined impact on CIMT in pediatric NS patients receiving ACEI or ARB therapy has not been extensively studied. Elucidating these relationships is essential for identifying potential therapeutic targets and guiding cardiovascular risk reduction strategies in this vulnerable population. Therefore, this study aims to evaluate the correlation between serum aldosterone and IL-6 levels with CIMT in children with SSNS and SRNS, before and after treatment with ACEI and ARB.

## MATERIALS AND METHODS

### Study settings and population

This prospective, single-arm pretest–posttest interventional study was conducted in the Pediatric Outpatient Unit of Dr. Zainoel Abidin Hospital, Banda Aceh, Indonesia. Baseline (pretest) assessments were conducted during May–June 2024, and follow-up (posttest) assessments were conducted from 24 August to 11 October 2024. The study evaluated changes in carotid intima-media thickness (CIMT), serum aldosterone, and interleukin-6 (IL-6) following treatment with lisinopril and valsartan. Participants were recruited consecutively, and the planned sample size was 30 participants. The trial was retrospectively registered with the Thai Clinical Trials Registry (TCTR20260923002). The trial was not registered before enrolment began because registration had not been completed at that time. The registration was completed on 23 September 2026. The authors acknowledge that retrospective registration limits prospective verification of the prespecified methods and outcomes. The authors confirm that all ongoing and related trials for this drug/intervention are registered.

This study enrolled children aged 2 to 18 years diagnosed with idiopathic NS and CKD based on established clinical guidelines [6]. Baseline (pretest) assessments were conducted during May–June 2024, and follow-up (posttest) assessments were conducted from 24 August to 11 October 2024. Idiopathic NS was defined by nephrotic-range proteinuria, hypoalbuminemia, and edema in the absence of secondary causes [6]. CKD was diagnosed as kidney damage or reduced glomerular filtration rate (GFR <60 mL/min/1.73 m^2^) persisting for more than three months [18]. Eligible patients had received steroid or other immunosuppressive therapy for at least six months and were classified as SSNS or SRNS. Patients must not have used ACEIs or ARBs within the previous month and were required to provide informed consent. Patients with infection, hypertensive urgency or emergency, stage 5 CKD requiring dialysis, or known allergy to ACEI or ARB preparations were excluded from the study. Patients who were lost to follow-up, withdrew consent during the study, or did not meet the minimum medication adherence were documented, and their data were handled according to predefined protocols.

### Types of participants

#### Inclusion criteria

Studies enrolling patients with CKD stages 1 to 4, as defined by the by Kidney Disease Outcomes Quality Initiative (K-DOQI) guidelines and who had albuminuria or proteinuria were considered for inclusion. We included studies in adults who had CKD regardless of etiology including individuals with diabetes complicated by kidney disease. The K/DOQI categories for kidney disease are:

- CKD stage 1: GFR > 90 mL/min/1.73 m^2^ and evidence of clinically relevant structural or urinary abnormalities including hematuria or proteinuria (or both)
- CKD stage 2: GFR 60 to 89 mL/min/1.73 m^2^
- CKD stage 3: GFR 30 to 59 mL/1.73 m^2^
- CKD stage 4: GFR 15 to 29 mL/min/1.73 m^2^

#### Exclusion criteria

We excluded studies in adults with CKD stage 5 (GFR < 15 mL/ min/1.73 m^2^) including those treated with dialysis and recipients of a kidney transplant.

Figure 1 illustrate flow diagram of participants through the study, from the enrollment to analysis.

**Figure 1.**
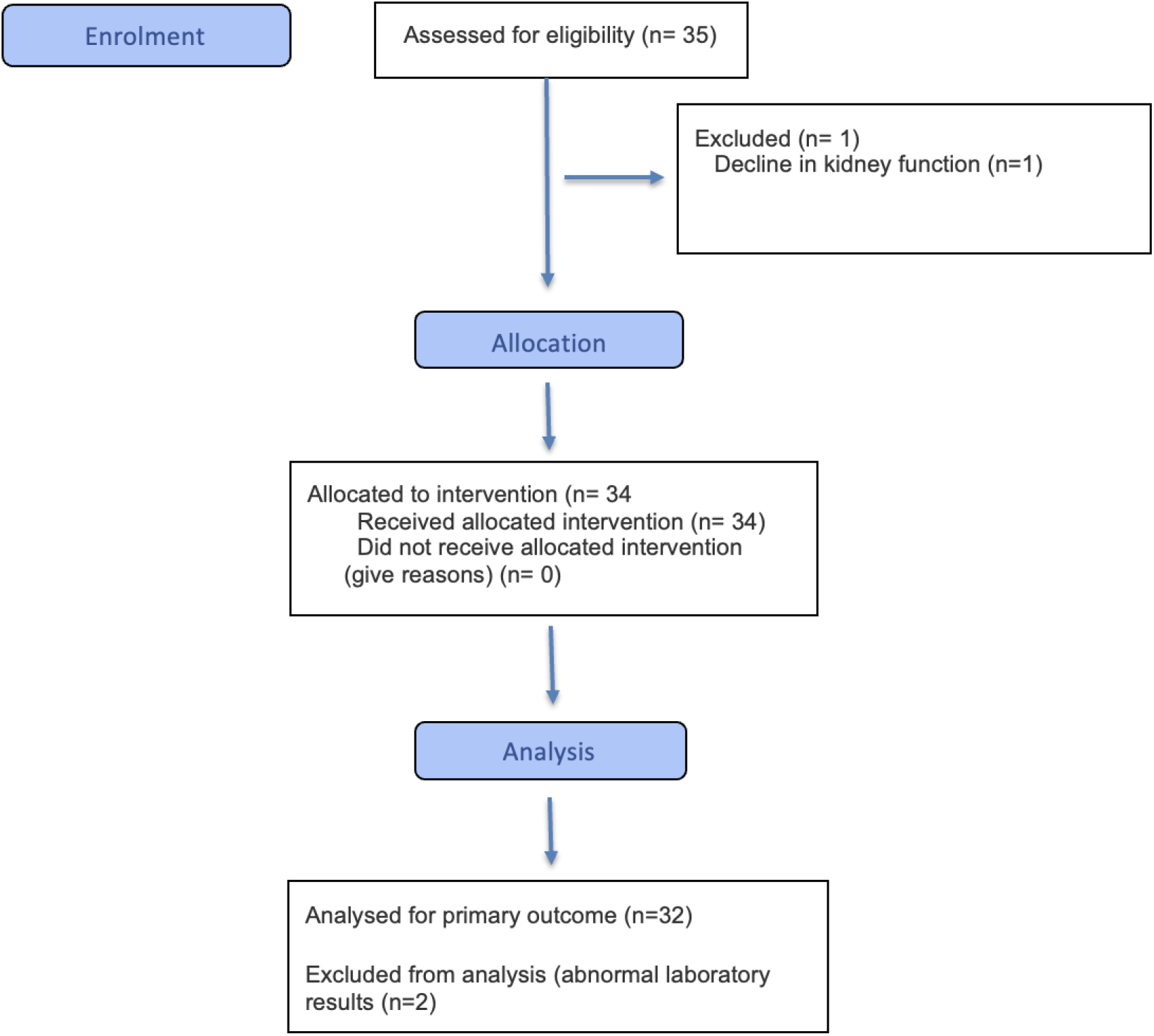
Consort Flow diagram of the participants

### Intervention

Patients were treated with lisinopril (ACEI) and valsartan (ARB) for 12 weeks. Doses were individualized based on body weight and age, and administered once daily in the evening. Medication adherence and potential adverse effects were monitored during clinic visits every four weeks, supplemented by biweekly telephone follow-ups. Patients and their caregivers received a logbook to record daily medication intake and any side effects, with caregivers acting as supervisors when possible. Treatment adjustments followed hospital protocols as necessary.

### Data Collection

Data were collected at baseline and after 12 weeks of treatment. Sociodemographic information (age, gender), clinical data (age at disease onset, NS classification, nutritional status, cyclosporine use, systolic and diastolic blood pressure), and laboratory parameters (complete blood count, total cholesterol, HDL cholesterol, LDL cholesterol, triglycerides, serum albumin, serum creatinine, and estimated glomerular filtration rate) were recorded. Urinalysis was also conducted.

To measure the serum aldosterone and IL-6 levels, venous blood samples (9 mL) were collected and divided into ethylenediaminetetraacetic acid (EDTA) tube for laboratory analysis. Serum aldosterone and IL-6 levels were measured using liquid chromatography– tandem mass spectrometry (LC-MS/MS) method (LC-MS/MS Waters Xevo TX-QS, Waters Corporation, Milford, MA, USA) and electrochemiluminescence immunoassay (Roche Diagnostics, Rotkreuz, Switzerland) respectively. The results are reported in ng/mL and pg/mL, respectively.

CIMT was assessed by a pediatric cardiologist using high-resolution B-mode Doppler ultrasound Philips iE33 (Philips Healthcare, Andover, MA, USA). Patients were positioned supine with the patient’s head elevated and rotated to optimize carotid artery imaging. CIMT measurements were performed using manual or automated edge detection methods and recorded in mm. Follow-up assessments during clinic visits monitored medication adherence, side effects, physical activity, and concomitant medications. Final blood sampling and CIMT measurements were repeated after the 12-week treatment. Patients with at least 80% medication adherence were included in the analysis. Protocol deviations were assessed by comparing the conduct of the study with the approved protocol and available study records. Any deviations identified, together with their timing and rationale, should be reported transparently.

### Statistical Analysis

All collected data were analyzed using SPSS version 26.0 (IBM Corp., Armonk, NY, USA). Descriptive statistics were applied to summarize patient characteristics and clinical parameters. Continuous variables, including serum aldosterone (ng/mL), serum IL-6 (pg/mL), and CIMT (mm), were assessed for normality using the Shapiro-Wilk test. Normally distributed data were presented as means ± standard deviations (SD), while non-normally distributed data were reported as medians and interquartile ranges. Correlation analysis between serum aldosterone, IL-6, and CIMT was conducted using either Pearson correlation tests based on the distribution of the variables. A *p*-value of <0.05 was considered statistically significant.

### Ethical approval

Ethics approval was obtained from the Research Ethics Committee, Dr. Zainoel Abidin Hospital (188/ETIK-RSUDZA/2024). Written informed consent was obtained from the parents or legal guardians of the selected participants. Data used in this study are available from the corresponding author upon reasonable request.

## RESULTS

Based on the initial data, the number of subjects who underwent the intervention was 35 individuals. However, during the course of the study, one subject was dropped out due to progressive decline in kidney function leading to death. Therefore, 34 subjects remained and were analyzed to assess laboratory parameters before and after the intervention. Furthermore, among these 34 subjects, abnormal laboratory results were found in two subjects compared to others, so they were excluded, resulting in a total of 32 subjects analyzed, as illustrated in the research flow diagram in Figure 1.

The subjects in this study were children with nephrotic syndrome aged between 2 and <18 years, totaling 32 children. This study aimed to determine the relationship between serum aldosterone levels and IL-6 with carotid artery intima-media thickness in patients with idiopathic nephrotic syndrome, in order to prevent cardiovascular abnormalities and progression to end-stage renal disease. The baseline characteristics of the study subjects are presented in Table 1.

**Table 1.** Characteristics of Respondent.

| Characteristics | SSNS<br>n = 15 | SRNS<br>n = 17 | Total<br>n = 32 | p |
| --- | --- | --- | --- | --- |
| Age (years old)* | 9.75 ± 4.93 | 10.61 ± 2.52 | 10.2 ± 3.8 | 0.551 |
| Sex, n (%) |  |  |  | 0.522 |
| Male | 8 (40) | 12 (60) | 20 (62.5) |  |
| Female | 7 (58.3) | 5 (41.7) | 12 (37.5) |  |
| Sistolik Blood Pressure (mmHg)* | 103.73 ± 13.01 | 105.76 ± 9.78 | 104.81 ± 11.27 | 0.619 |
| Diastolic Blood Pressure (mmHg)* | 66.26 ± 6 | 69.05 ± 10.95 | 67.75 ± 8.95 | 0.372 |
| Profile lipid serum** |  |  |  |  |
| Kolesterol (mg/dL) | 212 (145 - 393) | 211 (117 - 472) | 212 (117 - 472) | 0.762 |
| Trigliserida (mg/dL) | 121.6 (53 - 221) | 134 (56 - 231) | 128 (53 - 231) | 0.584 |
| HDL (mg/dL) | 46 (28 - 104) | 43 (27 - 126) | 46 (27 - 126) | 0.362 |
| LDL (mg/dL) | 151 (80 - 411) | 152 (69 - 411) | 152 (69 - 411) | 0.362 |
| Albumin (gr/dL)** | 3.2 (1.27 - 4.66) | 4.2 (1.86 - 4.8) | 3.72 (1.27 - 4.8) | 0.079 |
| Proteinuria, n (%) |  |  |  | 0.888 |
| Negative | 10 (47.6) | 11 (52.4) | 21 (65.6) |  |
| +1 | 2 (66.7) | 1 (33.3) | 3 (9.4) |  |
| +2 | 2 (28.6) | 5 (71.4) | 7 (21.9) |  |
| +3 | 1 (100) | 0 | 1 (3.1) |  |

Table 1 shows the baseline characteristics of thirty-two pediatric subjects with idiopathic nephrotic syndrome, divided into two groups: steroid-sensitive nephrotic syndrome (SSNS, n = 15) and steroid-resistant nephrotic syndrome (SRNS, n = 17). The mean age in the SSNS group was 9.75 ± 4.93 years, while in the SRNS group it was 10.61 ± 2.52 years, with an overall mean of 10.2 ± 3.8 years (p = 0.551), indicating no statistically significant age difference. In terms of sex distribution, the SSNS group consisted of eight boys (40%) and seven girls (58.3%), while the SRNS group consisted of twelve boys (60%) and five girls (41.7%). Overall, there were twenty boys (62.5%) and twelve girls (37.5%). The mean systolic blood pressure in the SSNS group was 103.73 ± 13.01 mmHg and in the SRNS group 105.76 ± 9.78 mmHg, with an overall mean of 104.81 ± 11.27 mmHg (p = 0.619), indicating no significant difference. The mean diastolic blood pressure was 66.26 ± 6 mmHg in the SSNS group and 69.05 ± 10.95 mmHg in the SRNS group, with an overall mean of 67.75 ± 8.95 mmHg (p = 0.372), also showing no significant difference.

The serum lipid profile is presented as median and range. Total cholesterol in the SSNS group was 212 mg/dL (145–393) and in the SRNS group 211 mg/dL (117–472), with an overall median of 212 mg/dL (117–472) (p = 0.762), indicating similar levels between groups. This aligns with reports that hypercholesterolemia is a common characteristic of nephrotic syndrome, both steroid-sensitive and resistant. Median triglyceride levels were 121.6 mg/dL (53–221) in the SSNS group and 134 mg/dL (56–231) in the SRNS group, with an overall median of 128 mg/dL (53–231) (p = 0.584), consistent with typical nephrotic syndrome hyperlipidemia due to increased lipoprotein synthesis and altered lipid metabolism. Median HDL levels were 46 mg/dL (28–104) in SSNS and 43 mg/dL (27–126) in SRNS, with an overall median of 46 mg/dL (27–127) (p = 0.362). Median LDL levels were 151 mg/dL (81– 440) in SSNS and 152 mg/dL (69–411) in SRNS, with an overall median of 152 mg/dL (69– 411) (p = 0.361). These findings are consistent with the pathophysiology of nephrotic syndrome, where total cholesterol and LDL tend to increase, while HDL may be normal or decreased due to impaired VLDL conversion and urinary loss of HDL.

Serum albumin levels in Table 1 are presented as median values: 3.2 g/dL (1.27–4.66) in the SSNS group and 4.2 g/dL (1.86–4.8) in the SRNS group, with an overall median of 3.72 g/dL (1.27–4.8) (p = 0.079). This reflects hypoalbuminemia, which is commonly observed in nephrotic syndrome due to protein loss through urine and contributes to edema and lipid metabolism alterations. Proteinuria was assessed semi-quantitatively (negative, +1, +2, +3). In the SSNS group, ten children (47.6%) were negative, two children (66.7% of positives) had +1, two children (28.6%) had +2, and one child (100%) had +3. In the SRNS group, eleven children (52.4%) were negative, one child (33.3%) had +1, five children (71.4%) had +2, and none had +3. Overall, 65.6% of subjects were in the negative category (p = 0.888), indicating no significant difference in proteinuria distribution between groups, although clinically +1 to +3 still reflect increased protein leakage and glomerular damage.

Figure 2 presents a bar diagram illustrating changes in median serum urea, creatinine, and potassium levels before and after ACEI-ARB therapy in patients with idiopathic nephrotic syndrome. Median serum urea decreased from approximately 30–35 mg/dL before therapy to about 23.6 mg/dL after therapy. Serum creatinine decreased from 0.70 mg/dL to 0.429 mg/dL after therapy. Potassium levels increased from a median of 3.8 mmol/L before therapy to 4.2 mmol/L after therapy. Overall, the diagram illustrates improvement in kidney function (decreased urea and creatinine) accompanied by a controlled increase in potassium levels. Figure 2. Analysis of kidney profile differences before and after ACEI-ARB therapy administration

**Figure 2.**
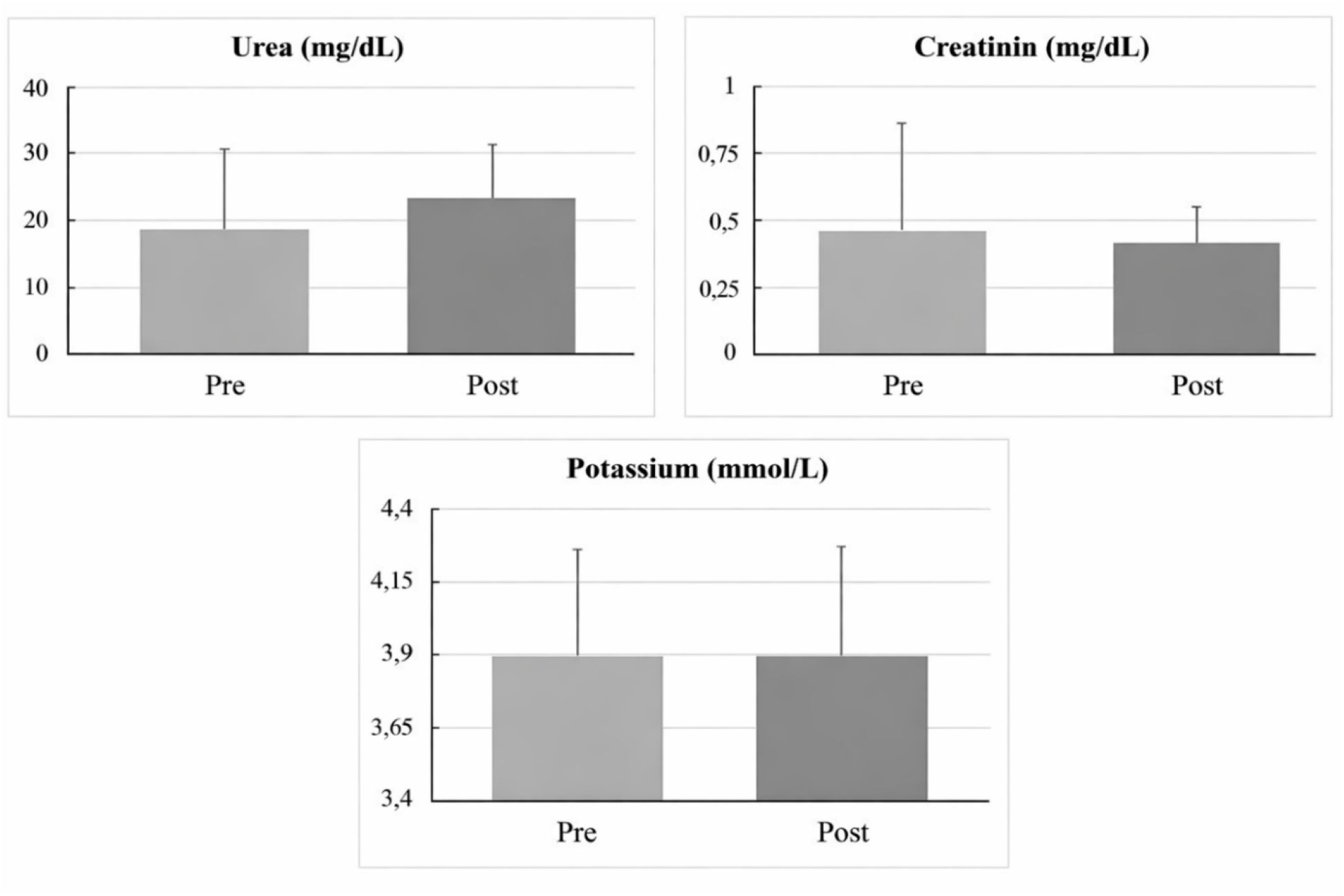
Analysis of kidney profile differences before and after ACEI-ARB therapy administration

Table 2 presents an analysis of differences in serum aldosterone levels before and after ACEI and ARB therapy in thirty-two pediatric subjects with idiopathic nephrotic syndrome. The mean aldosterone level before therapy was 5.15 ng/dL and after therapy 4.0 ng/dL, with p = 0.933 (not significant). This indicates that RAAS blockade with ACEI and ARB did not result in a significant reduction in serum aldosterone levels, although theoretically these agents reduce angiotensin II production, which stimulates aldosterone secretion from the adrenal zona glomerulosa. This phenomenon is explained by “aldosterone escape,” where aldosterone levels remain stable or increase despite prolonged RAAS blockade, as reported in studies of chronic kidney disease patients. Overall, aldosterone levels remained within the normal physiological range for children, with a decreased min–max range after therapy, although not statistically significant.

**Table 2.** Analysis of differences in serum aldosterone levels before and after ACEI-ARB therapy administration.

| Aldosterone (ng/dL) | Median (min - max) | p* |
| --- | --- | --- |
| Pre, n = 32 | 5.15 (0.6 - 49.32) | 0.933 |
| Post, n = 32 | 4.05 (0.6 - 70.48) |  |

\* Wilcoxon rank test

Table 3. analyzes serum IL-6 levels before and after therapy. The mean IL-6 level was 3.5 pg/mL both before and after therapy, with p = 0.156 (not significant, p > 0.05). This indicates that ACEI/ARB therapy did not affect serum interleukin-6 levels, a pro-inflammatory cytokine involved in renal inflammation and fibrosis in nephrotic syndrome. IL-6 is often elevated in kidney diseases such as glomerulonephritis and diabetic nephropathy; however, in this study, baseline levels were already low (around 3.5 pg/mL), within the normal range (<7 pg/mL in healthy children), which limited the reduction potential. The Wilcoxon rank test (p = 0.156) further confirmed no significant difference in IL-6 levels before and after ACEI-ARB therapy.

**Table 3.** Analysis of differences in serum IL-6 levels before and after ACEI-ARB therapy administration.

| IL-6 (pg/mL) | Median (min - max) | p* |
| --- | --- | --- |
| Pre, n = 32 | 3.5 (3.5 - 27.1) | 0.156 |
| Post, n = 32 | 3.5 (3.5 - 9.64) |  |

\* Wilcoxon rank test

Based on Table 4, the carotid artery intima-media thickness (CIMT) before therapy had a mean of 0.13 ± 0.04 cm and decreased to 0.06 ± 0.01 cm after therapy, with a statistically significant p-value of <0.001. The percentage change ranged from 0.04 to 0.08 cm, indicating a meaningful reduction in intima-media thickness as a marker of subclinical atherosclerosis. The test results (p < 0.001) confirm the renoprotective and vasculoprotective effects of ACEI/ARB therapy in preventing vascular remodeling in nephrotic syndrome. The pre-therapy CIMT value of 0.13 cm reflects early cardiovascular risk due to hyperlipidemia and chronic inflammation in nephrotic conditions. The post-therapy decrease to 0.06 cm, with an overall negative mean percentage change, indicates a reversal of intima-media thickening. This effect is achieved through the reduction of oxidative stress, endothelial inflammation, and intravascular pressure via RAAS inhibition. Similar studies in children with idiopathic nephrotic syndrome have identified total cholesterol and steroid-resistant nephrotic syndrome (SRNS) status as predictors of CIMT, making this reduction relevant for long-term cardiovascular complication prevention.

**Table 4.** Analysis of CIMT differences before and after ACEI-ARB therapy administration.

| CIMT (cm) | Mean ± SD | p* | Mean Difference (95% CI) |
| --- | --- | --- | --- |
| Pre, n = 32 | 0.13 ± 0.04 | <0.001 | 0.06 (0.04 - 0.08) |
| Post, n = 32 | 0.06 ± 0.01 |  |  |

\* Paired sample t-test

Based on Figure 3, the scatter plot diagram illustrates a linear correlation between changes in serum aldosterone levels and changes in carotid intima-media thickness (CIMT) after ACEI-ARB therapy in pediatric subjects with idiopathic nephrotic syndrome. The linear regression line shows a moderate positive slope, with a very weak coefficient of determination (R^2^ = 0.075), indicating that only 7.5% of the variability in CIMT changes is explained by changes in aldosterone levels. Several right-sided outliers (e.g., aldosterone increase of +40 ng/dL) are associated with high positive CIMT changes (+1.5 mm), supporting the positive trend of the regression line despite high dispersion. This diagram contrasts with the previous multivariate regression table, where changes in aldosterone (B = 0.001; p = 0.184) were not statistically significant (confidence interval includes zero), consistent with the weak univariate R^2^ value of 0.075, emphasizing the minimal independent effect. Figure 3. Analysis of the correlation between aldosterone changes and CIMT changes after ACEI-ARB therapy administration.

**Figure 3.**
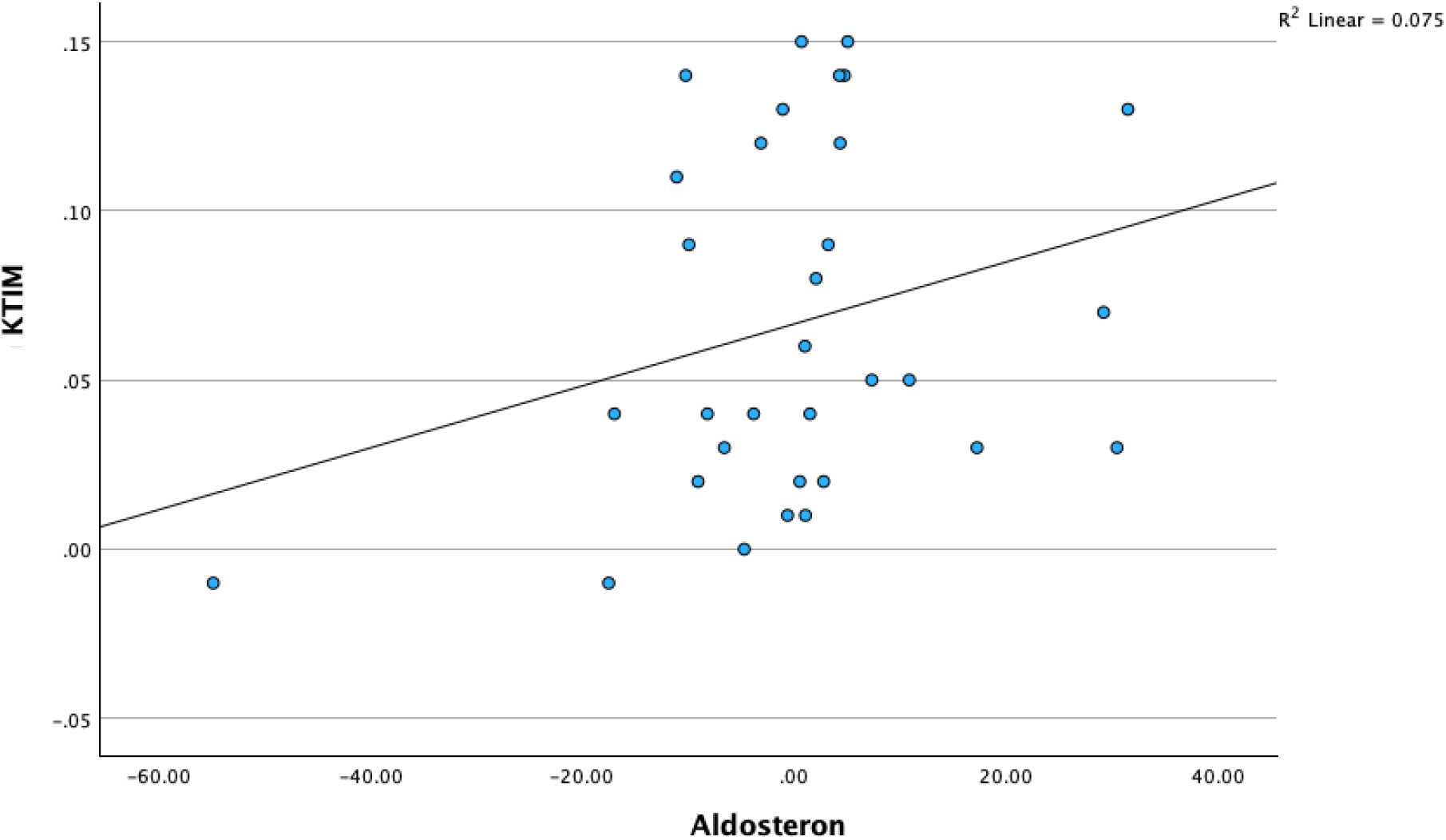
Analysis of the correlation between aldosterone changes and CIMT changes after ACEI-ARB therapy administration.

Based on Table 5, which presents the analysis of differences in serum aldosterone levels between steroid-sensitive nephrotic syndrome (SSNS) and steroid-resistant nephrotic syndrome (SRNS) groups, the aldosterone level in the SSNS group was 1.02 ng/dL, within the normal physiological range for children (0.5–20 ng/dL), reflecting stable hormone levels after renin-angiotensin-aldosterone system (RAAS) blockade, although ACEI/ARB theoretically suppresses aldosterone secretion by reducing angiotensin II. In the SRNS group, the median aldosterone level was 0.7 ng/dL. The Mann-Whitney U test yielded a p-value of 0.664, indicating no significant difference in serum aldosterone changes between the two types of nephrotic syndrome. The absence of this difference suggests that other factors, such as the type of nephrotic syndrome, do not mediate vascular remodeling through aldosterone.

**Table 5.** Analysis of differences in serum aldosterone changes based on nephrotic syndrome type.

| Aldosterone Changes | Median (min, max) | p* |
| --- | --- | --- |
| SSNS, n = 15 | 1.02 (-54.86, 29.23) | 0.664 |
| SRNS, n = 17 | 0.7 (-16.97, 31.52) |  |

\* Mann Whitney test, SSNS: steroid-sensitive nephrotic syndrome; SRNS: steroid-resistant nephrotic syndrome

The scatter plot diagram in Figure 4 illustrates the linear relationship between changes in serum IL-6 levels and changes in carotid intima-media thickness (CIMT) after ACEI-ARB therapy in children with idiopathic nephrotic syndrome. The regression line shows an upward positive slope of moderate gradient, with a coefficient of determination (R^2^ = 0.215), indicating that 21.5% of the variability in CIMT changes can be explained by changes in serum IL-6 levels. Figure 4. Analysis of the correlation between IL-6 changes and CIMT changes after ACEI-ARB therapy administration.

**Figure 4.**
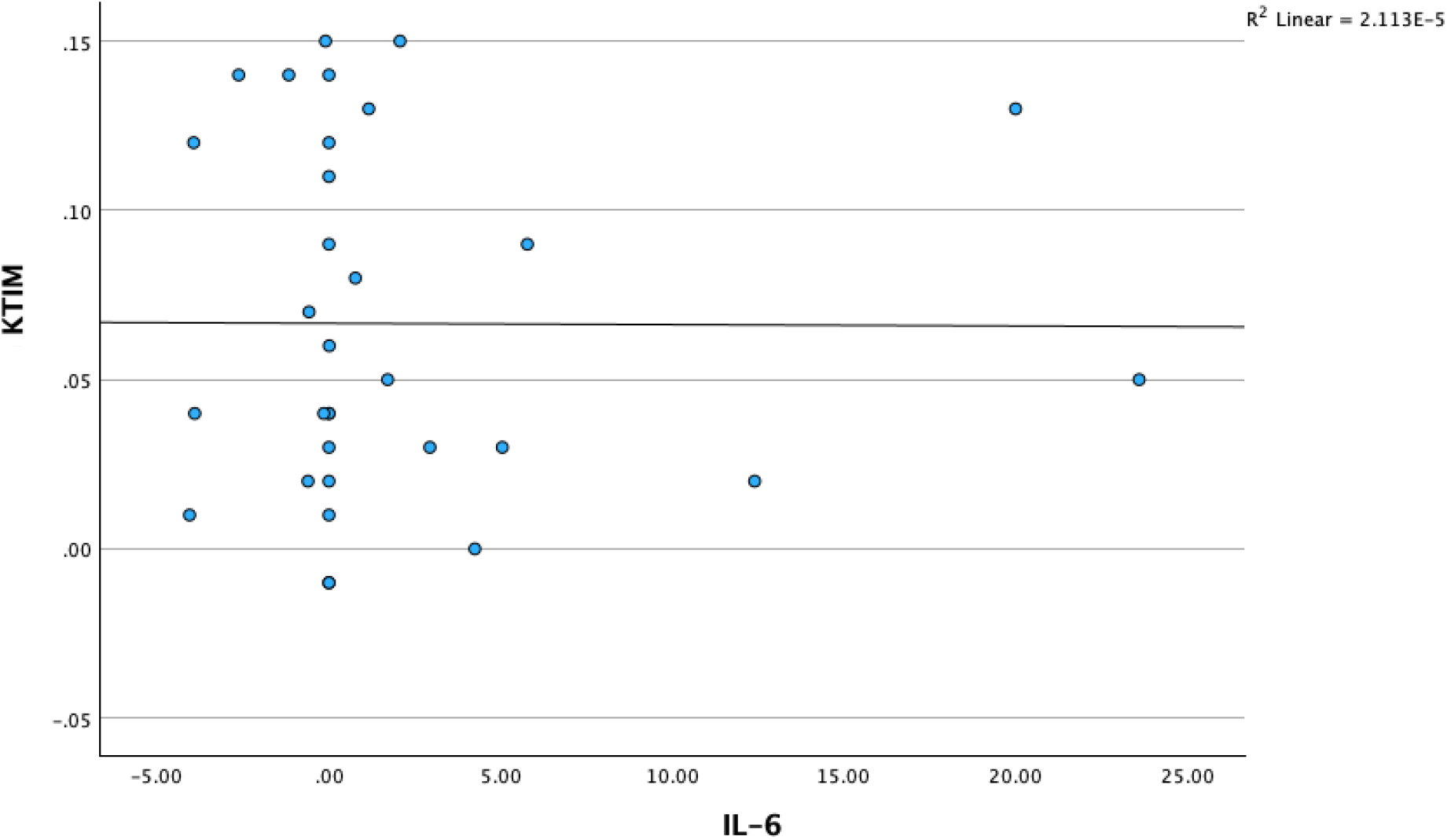
Analysis of the correlation between IL-6 changes and CIMT changes after ACEI-ARB therapy administration.

The data points are distributed with a clear positive trend, where negative changes in IL-6 (decreases, left of the x-axis from 0 to -6 pg/mL) are mostly correlated with negative changes in CIMT (reduction in vascular thickening, below the y-axis from -1 to 0 mm), suggesting that reduced inflammation contributes to the therapeutic benefit. Based on Table 6, which presents the analysis of differences in serum interleukin-6 (IL-6) levels between the steroid-sensitive nephrotic syndrome (SSNS, n = 15) and steroid-resistant nephrotic syndrome (SRNS, n = 17) groups using the Mann–Whitney test, the p-value of 0.476 indicates no statistically significant difference in IL-6 changes between the two groups after ACEI-ARB therapy.

**Table 6.** Analysis of differences in IL-6 changes based on nephrotic syndrome type.

| IL-6 Changes | Median (min, max) | p* |
| --- | --- | --- |
| SSNS, n = 15 | 0 (-4.06, 20) | 0.476 |
| SRNS, n = 17 | 0 (-3.94, 23.6) |  |

\* *Mann Whitney test*, SSNS: steroid-sensitive nephrotic syndrome; SRNS: steroid-resistant nephrotic syndrome

Multivariate analysis was performed to assess the relationship between risk factors and changes in carotid intima-media thickness (CIMT) in children with nephrotic syndrome. The results, as shown in Table 7, indicate that changes in IL-6 had a coefficient of B = -9.653 (SE = 0.002, p = 0.953, 95% CI: -0.003 to 0.003). A p-value greater than 0.05 indicates that IL-6 changes do not contribute to CIMT changes. Changes in aldosterone showed B = 0.001 (SE = 0.001, p = 0.184, CI: 0.00–0.002, β = 0.00%), indicating a minimal and non-significant positive effect. Systolic blood pressure (B = 0, SE = 0.001, p = 0.939) and diastolic blood pressure (B = 0.001, SE = 0.002, p = 0.418) did not predict vascular remodeling. Albumin levels (B = 0.002, SE = 0.01, p = 0.847) also showed no significant effect, indicating that hypoalbuminemia was not associated with CIMT changes. The type of nephrotic syndrome showed B = 0.02 (SE = 0.01, p = 0.047, CI: 0.00–0.04, β = -0.19%), indicating a statistically significant association (p < 0.05), where SRNS was positively associated with greater CIMT changes compared to SSNS. Sex showed B = -0.015 (SE = 0.01, p = 0.23, CI: -0.04 to 0.01, β = -0.09%), indicating no significant association.

## DISCUSSION

Carotid intima-media thickness (CIMT) has emerged as a reliable non-invasive marker for assessing subclinical atherosclerosis in pediatric patients with nephrotic syndrome (NS) [19–22]. It reflects early vascular remodeling and correlates with long-term cardiovascular risk. In the present study, a significant reduction in CIMT was observed following ACEI/ARB therapy in both steroid-sensitive nephrotic syndrome (SSNS) and steroid-resistant nephrotic syndrome (SRNS) groups. Specifically, CIMT decreased from 0.132 mm to 0.065 mm in SRNS and from 0.130 mm to 0.067 mm in SSNS. These findings are consistent with previous studies demonstrating that inhibition of the renin–angiotensin–aldosterone system (RAAS) attenuates vascular remodeling [8,23]. Therefore, CIMT may serve as a valuable surrogate endpoint for evaluating therapeutic efficacy in pediatric NS.

The greater reduction in CIMT observed in SRNS patients may be attributed to their higher baseline vascular burden. Chronic inflammation, persistent proteinuria, and prolonged hypertension contribute to more severe endothelial dysfunction in this group [13,14]. These factors enhance RAAS activation, which plays a central role in vascular injury and remodeling [9]. Consequently, RAAS inhibition may yield a more pronounced relative benefit in SRNS patients. This observation suggests that even patients with more severe disease can achieve meaningful vascular improvement. It also underscores the importance of early cardiovascular risk assessment in high-risk populations [24].

Despite the significant reduction in CIMT, no statistically significant correlations were identified between aldosterone or IL-6 levels and CIMT changes. This finding suggests that these biomarkers may influence cardiovascular risk through indirect or non-structural pathways [16,25]. Aldosterone and IL-6 are known mediators of inflammation and fibrosis; however, their effects may not immediately translate into measurable vascular structural changes. Vascular remodeling is a complex and multifactorial process involving hemodynamic, metabolic, and inflammatory components [26]. Therefore, reliance on single biomarkers may not adequately reflect overall cardiovascular risk. These findings highlight the need for a more integrative approach to risk stratification in NS.

The paradoxical increase in aldosterone levels following ACEI/ARB therapy observed in this study is consistent with the phenomenon known as “aldosterone escape” [12,27]. This condition is characterized by persistent or increased aldosterone levels despite effective RAAS blockade. Several mechanisms have been proposed, including alternative pathways for aldosterone synthesis and stimulation by potassium or adrenocorticotropic hormone [18]. Aldosterone escape may limit the long-term effectiveness of ACEI/ARB therapy in suppressing RAAS activity. Consequently, this phenomenon provides a rationale for considering adjunctive therapies such as mineralocorticoid receptor antagonists. However, further studies are needed to clarify its clinical significance in pediatric populations.

Aldosterone contributes to vascular and renal injury through activation of mineralocorticoid receptors, leading to oxidative stress, inflammation, and extracellular matrix deposition [18]. These processes promote both vascular remodeling and renal fibrosis. Experimental and clinical evidence suggests that aldosterone exerts effects independent of angiotensin II, further supporting its role in disease progression [12]. Incomplete suppression of aldosterone during RAAS inhibition may therefore perpetuate ongoing tissue damage. The addition of aldosterone antagonists has been shown to further reduce proteinuria [27]. However, the benefits must be carefully weighed against potential risks. One of the major limitations of aldosterone antagonist therapy is the increased risk of hyperkalemia, particularly in patients with impaired renal function [28]. Reduced potassium excretion in CKD predisposes patients to electrolyte imbalances. Studies have demonstrated that combining RAAS inhibitors with aldosterone antagonists significantly increases this risk [27]. Additionally, adverse effects such as gynecomastia have been reported, especially with non-selective agents like spironolactone. These safety concerns necessitate careful patient selection and close biochemical monitoring. Therefore, individualized treatment strategies are essential when considering combination therapy [12].

The improvement in renal function parameters observed in this study further supports the renoprotective effects of ACEI/ARB therapy. Reductions in serum urea and creatinine indicate improved glomerular hemodynamics. This effect is primarily mediated by decreased intraglomerular pressure and reduced protein filtration [8]. Lower proteinuria not only reflects improved renal function but also reduces tubular injury and inflammation. Consequently, the progression of chronic kidney disease may be slowed. These findings reinforce the central role of RAAS inhibition in clinical nephrology [1,5].

Interleukin-6 (IL-6) is a pro-inflammatory cytokine that plays a key role in endothelial dysfunction and vascular injury [25,29]. It contributes to oxidative stress, leukocyte adhesion, and reduced nitric oxide bioavailability. However, IL-6 levels did not change significantly following ACEI/ARB therapy in this study. This may be explained by relatively low baseline IL-6 levels or the short duration of follow-up. Additionally, inflammatory responses in NS involve multiple cytokines, not solely IL-6. Therefore, IL-6 alone may not adequately represent the inflammatory burden in these patients.[17] The differential changes in IL-6 between SRNS and SSNS groups provide further insight into disease mechanisms. A reduction in IL-6 was observed in SRNS, suggesting a potential anti-inflammatory effect of therapy in more severe disease [30]. However, the lack of correlation with CIMT indicates that inflammation may not directly translate into structural vascular changes in the short term. Other factors such as dyslipidemia, hypertension, and oxidative stress may play a more dominant role. This highlights the multifactorial nature of cardiovascular risk in NS. Comprehensive management strategies are therefore required [22,31].

Several methodological limitations should be considered when interpreting the findings of this study. The relatively small sample size and short follow-up period may limit the generalizability of the results. Most available studies, including the present one, rely on surrogate endpoints such as CIMT and proteinuria [22]. Data on hard clinical outcomes, including cardiovascular events and progression to end-stage kidney disease, remain limited. Furthermore, heterogeneity in patient characteristics and treatment regimens may influence outcomes. Larger and longer-term studies are needed to confirm these findings [28].

The clinical implications of this study are important for the management of pediatric NS. Routine CIMT assessment may be considered for early detection of subclinical vascular changes, particularly in high-risk patients such as those with SRNS [20]. ACEI/ARB therapy should remain a cornerstone of treatment, even in steroid-resistant cases [6,7]. However, clinicians should remain vigilant regarding potential adverse effects, especially when considering combination therapy. Regular monitoring of renal function and electrolyte levels is essential. Personalized treatment approaches based on individual risk profiles are strongly recommended. [11,12,18].

## CONCLUSION

This study highlights the importance of comprehensive cardiovascular risk management in pediatric nephrotic syndrome. While RAAS inhibition significantly improves vascular and renal parameters, it does not fully address all underlying pathogenic mechanisms. Aldosterone escape and persistent inflammatory pathways remain important therapeutic challenges. Future research should focus on identifying additional therapeutic targets and optimizing combination strategies. Long-term studies are required to evaluate the impact on clinical outcomes beyond surrogate markers. Ultimately, an integrated and individualized approach will be essential to improve long-term prognosis.

## Data Availability

Thank You

## Acknowledgements

We would like to thank all participants involved in this study

## Acknowledgments

The content of this publication does not necessarily reflect the views or policies of the Department of Health and Human Services, nor does mention of trade names, commercial products, or organizations.

